# Gene-Set Based Rare Variant Association Analysis of Whole Genome Sequencing Data in the Portuguese Island Collection for Schizophrenia and Bipolar Disorder

**DOI:** 10.64898/2026.05.28.26354351

**Authors:** Hamed Kazemi, John Drake, Tim Bigdeli, Silviu Bacanu, Tan Hoang Nguyen, Kelly Benke, Brion Maher, James Knowles, Steve McCarroll, Celia Carvalho, Helena Medeiros, Rute Ferreira, Michele Pato, Carlos Pato, Vladimir Vladimirov, Ayman Fanous

## Abstract

Schizophrenia (SCZ) and bipolar disorder (BPD) are highly heritable psychiatric disorders with complex polygenic architectures. Genome-wide association studies (GWASs) have identified numerous common variant associations, but rarer variants detectable through whole-genome sequencing (WGS) remain underexplored. We conducted rare variant association analysis using WGS data from the Portuguese Island Collection (PIC), including 28 families with SCZ (n = 53) and 41 families with BPD (n = 83) cases, and population controls (n = 62).

Following ANNOVAR and CADD annotation, burden analysis of deleterious variants showed that both affected and unaffected family members from SCZ and BPD pedigrees had significantly higher burdens of rare deleterious variants compared to controls (p < 0.0001), with no significant differences observed between affected and unaffected relatives, consistent with shared familial genetic liability. Polygenic Risk Score (PRS) analysis confirmed significant genetic contributions to both disorders within PIC. Association analyses were subsequently performed using SAIGE-GENE+ identifying 483 and 583 nominally significant (suggestive associations) gene sets (p-value ≤ 0.05; FDR > 0.05) for SCZ and BPD, respectively, including gene sets related to neurotransmission, synaptic function and structure, neurodevelopment, and neuroinflammation as well as major signaling pathways. Cross disorder overlaps also identified shared suggestive enrichment of GABA and glutamate signaling, synaptic signaling, and Wnt signaling gene sets in both SCZ and BPD.

These findings support shared rare variant burden within multiplex psychiatric families and highlight the role of gene-set based rare variant analysis in identifying neurobiological pathways relevant to SCZ and BPD.

## Introduction

Schizophrenia (SCZ) and Bipolar Disorder (BPD) rank among the leading 20 contributors to global disability, each affecting approximately 1% of the general population (1). While the etiology of these disorders remains largely unclear, it is well-established that genetics plays a pivotal role, with heritability estimates ranging from 60% to 80% for SCZ and 57% to 85% for BPD (2–5). There is mounting evidence that the genetic architecture of SCZ and BPD is polygenic and encompasses both common and rare genetic variants, which are found across the genome including noncoding regions (1, 6). Large-scale GWASs, such as the recent Psychiatric Genomics Consortium (PGC) schizophrenia GWAS (7), have identified common risk loci associated with SCZ, implicating synaptic and neuronal processes in disease pathology (8). This diversity in genetic contribution has been conceptualized through models that aim to explain how different variants affect disease susceptibility. Although there has been ongoing debate over common and rare variants, the prevailing view is that both common (low penetrance) and rare (high penetrance) variants contribute to genetic risk for common diseases, with each type potentially influencing disease susceptibility in different ways (9). While common variants account for a portion of the heritability of genetic disorders, a substantial gap known as missing heritability remains in explaining the genetic risk (10, 11). This missing heritability has led to increasing recognition of the contribution of rare genetic variants.

The rare variants are thought to contribute to a portion of the unexplained heritability of SCZ and BPD (11, 12), particularly within multiplex families which may be enriched for inherited deleterious variation (13, 14), further highlighting the complex genetic heterogeneity underlying these neuropsychiatric disorders (13, 15). Rare coding genetic variants identified via whole-exome sequencing (WES) studies have been implicated in the etiology of SCZ and BPD (14, 16, 17), particularly ultra-rare variants (URVs; minor allele frequency (MAF) ≤ 0.001) that may disrupt protein-coding gene regulation (7, 18). Also, the rare variants alter noncoding RNA gene exons or splicing and transcription regulatory motifs, which are typically located outside of protein-coding exon (15). Consequently, WGS is necessary to comprehensively evaluate both coding and noncoding rare variation. By enabling genome-wide interrogation at single nucleotide resolution, WGS can broaden the detection of rare variants, improves gene-based analyses, and provides more complete coverage of genomic regions compared to WES (12, 19, 20).

Findings from case-control and family-based studies indicate overlapping genetic risk factors for SCZ and BPD, with a genetic correlation of around 0.7 (4, 21, 22). This correlation has prompted further investigations into the specific genetic loci that may contribute to both disorders, including linkage analyses in PIC families that identified overlapping chromosomal regions associated with SCZ and BPD (23), as well as recent cross-ancestry multivariate GWAS demonstrating extensive shared genetic architecture, particularly between SCZ and BPD (24). Family-based studies provide more statistical power for identifying rare variants since families with multiple affected members are more likely to share disease-specific variants (25). Previous WGS studies in multiplex families have identified rare and deleterious variants in genes involved in neurodevelopmental and synaptic pathways (13, 18, 26–28). For instance, deleterious variants in brain-expressed genes co-segregated with SCZ in several pedigrees (25), and a higher prevalence of rare and deleterious variants in neuron projection genes in SCZ patients with 22q11.2 deletions (15). The Schizophrenia Exome Meta-Analysis (SCHEMA) study also identified highly deleterious URVs in key SCZ risk genes that influence synaptic structure and function, neuronal migration, transcriptional regulation, and ubiquitin ligation (7). The ultra-rare pathogenic variants in genes involved in the glutamatergic neurotransmission pathway, were also found to segregate in SCZ multiplex families (29). In BPD, genetic risk variants have been associated with structural alterations in the brain (30). Moreover, family-based WGS/WES studies identified rare pathogenic variants in genes associated with presynaptic neurotransmission and postsynaptic density regulation, suggesting the potential contribution both synaptic compartments in SCZ and BPD (31, 32). Furthermore, the significant genetic heterogeneity in SCZ and BPD impedes the identification of rare alleles (1, 33). To address this challenge, a homogeneous or isolate population, such as the Portuguese Island Collection (PIC) from the Portuguese islands, including the Azores and Madeira, offers improved power by enriching rare variants through genetic drift (1). This population has been useful for linkage studies (23) and has contributed to the large-scale PGC efforts.

Analytical approaches such as the Scalable and Accurate Implementation of GEneralized mixed model (SAIGE) and its extension, SAIGE-GENE+ (34–36) as well as gene-set based analyses (37) have proven invaluable tools for investigating the segregation of rare variants derived from WGS data and their association with neuropsychiatric disorders. SAIGE is based on a generalized linear mixed model framework, enabling effective control for population structure and sample relatedness while also addressing case–control imbalance and efficiently improving power for rare variant association testing (34–36, 38, 39). SAIGE-GENE+ extends this framework to set-based rare variant testing by aggregating variants within genes or regions, improving calibration and efficiency under case–control imbalance, and enabling flexible grouping by functional annotation and allele frequency to capture cumulative effects (36). Building on this, gene-set analysis offers insights into the cumulative impact of rare variants across genes within biological pathways especially those relevant to the neuropathology of psychiatric disorders, providing a comprehensive understanding of the genetic architecture and etiology of SCZ and BPD (40, 41). Here, we present our analyses, including Polygenic Risk Score (PRS), permutation-based validation, burden analyses of deleterious variants, and gene-set based rare variant association test on WGS data using SAIGE-GENE+ in PIC families segregating for SCZ and BPD.

## Materials and methods

### Recruitment and phenotyping for Portuguese Island Collection (PIC)

This study was designed to assess the association of rare variants through gene set-based method with SCZ and BPD risk in families collected as part of the PIC, which were ascertained in the Azores, Madeira, and Mainland Portugal. It includes affected and unaffected family members and nonfamilial population controls. The rationale for using this population is to reduce genetic heterogeneity by drawing from a homogenous population relative to outbred populations, thus increasing the power to detect disease predisposing variants (42–44). Indeed, the PIC sample was shown to be more homogenous, with the Azorean population demonstrating the strongest evidence for homogeneity (45). As previously described (46), blind best-estimate diagnoses were made using a Portuguese translation of the Diagnostic Interview for Genetic Studies. Our initial dataset consisted of 359 individuals including SCZ cases (n = 59), BPD cases (n = 87), unaffected individuals (n = 110), family members with other psychiatric disorders (n = 41), and nonfamilial population controls with no family history of psychiatric disorders (n = 62). The overall sex distribution of the cohort was 155 males and 204 females. Geographically, 208 individuals were from the Azores, 112 from Madeira, and 39 from Mainland Portugal (Supplementary File 1).

### WGS data

Genomic DNA (350 ng in 50 µL), isolated from the whole blood, was sheared to approximately 385 bp using a Covaris focused-ultrasonicator, followed by solid phase reversible immobilization (SPRI) size selection. Libraries were prepared with the KAPA Hyper Prep kit (KAPA Biosystems) and indexed using Roche adapters. Quantification was performed via qPCR, and libraries were normalized to 2.2 nM and pooled. Cluster amplification and sequencing were conducted on the Illumina HiSeq X and NovaSeq 6000 platforms, producing 151 bp paired-end reads. The resulting data were processed using the Picard pipeline and aligned to the GRCh38 reference genome using BWA-MEM. Outputs were generated as demultiplexed CRAM or BAM files, with sample tracking managed by an automated LIMS (Broad Institute).

### Quality Control

Plink 1.9 (47) was used to perform a sex check, accounting for the pseudo-autosomal regions, and check for excessive Mendelian errors and low-quality reads. After excluding problematic subjects (e.g., F coefficient > 0.3 for females and < 0.7 for males), and subjects diagnosed with psychiatric disorders other than SCZ or BPD, the final dataset comprised 300 individuals including nonfamilial population controls (n = 62), unaffected individuals with family history of SCZ (n = 66), affected individuals with SCZ (n = 53), unaffected individuals with family history of BPD (n = 63), and affected individuals with BPD (n = 83) (Supplementary File 2). In total, there were 41 families with affected BPD individuals and 28 families with affected SCZ individuals. Notably, 27 unaffected individuals were shared between the unaffected BPD and unaffected SCZ groups. Prior to quality control filtering, the dataset contained 31,445,428 variants. Rare autosomal variants were validated using Genome Aggregation Database (gnomAD) v4 non-Finnish European population. Rare variants were defined as autosomal variants with sufficient coverage (call rate ≥ 75%) and a MAF between 1×10⁻⁶ and 0.01. In total, 8,550,822 validated rare variants were identified and extracted for further analysis.

### Calculation of PRS and Multidimensional scaling (MDS) analysis of 1000 genome with PIC

Calculation of PRS was carried out using PRSice 2.3.3 (48). We used PIC data as the target dataset and the latest GWAS summary statistics for SCZ (8) and BPD (49) from PGC website (https://pgc.unc.edu/for-researchers/download-results/) as the base dataset. R 4.1.3 (https://cran.r-project.org/bin/windows/base/old/4.1.3/) and Plink 1.9 were used for quality control for the base and target data. We performed standard quality control for base and target datasets, checking for sex, mismatching, duplicate, and ambiguous single nucleotide polymorphisms (SNPs). We considered two different analyses: a) SCZ/BPD cases versus unaffected family members, and b) SCZ/BPD cases versus population controls. The first analysis assesses whether PRS can distinguish cases from genetically similar individuals, thereby capturing a within-family signal. The second analysis evaluates differences in polygenic burden between cases and unrelated individuals, reflecting a between-family signal. In addition, we conducted MDS analysis using the 1000 genome dataset (build 38, 3202 samples) (50) to investigate the genetic relationships and population structure of the PIC population relative to the Iberian-Spanish population. Specifically, we sought to determine whether some families in the PIC population cluster separately. Plink v1.9 was used for MDS analysis. Both the PIC and 1000 genome datasets were filtered for matching variants and subsequently merged into a single dataset comprising 3,718,966 variants. Filtering parameters included MAF ≤ 0.05 and linkage disequilibrium (LD) pruning. For MDS analysis, pairwise Identity-by-State (IBS) distances and the first 20 dimensions of the MDS plot were used as parameters. The result of the MDS analysis is provided in the supplementary documents (Supplementary File 3).

### Annotation of variants

Deleterious variants were annotated using two metrics. Variants were functionally annotated using ANNOVAR (v2020.06.08) (51) and the GRCh38 reference genome with refGene database. Then, Combined Annotation Dependent Depletion (CADD) v1.6 (52) (https://cadd.bihealth.org/) with GRCh38 was used to assign PHRED-like scores to variants. CADD is a scaled metric that ranks the deleteriousness of SNPs and small indels across the genome by integrating diverse functional annotations into a single score, and higher scores indicate greater predicted deleteriousness. We focused on variants in the top 10% (e.g., CADD-PHRED score ≥ 10). In total, 37,507 and 451,353 rare deleterious variants were identified by ANNOVAR and CADD, respectively.

### Burden analysis of deleterious variants

To assess differences in deleterious variant burden among affected individuals, unaffected relatives, and population controls, linear mixed-effects models were fitted separately for SCZ and BPD cohorts using deleterious variants defined by both ANNOVAR annotation and CADD-PHRED scores. Individual-level burden was calculated as the total number of deleterious variants carried by each subject. Because individuals within multiplex pedigrees are not statistically independent, family relationship was incorporated as a random effect in the model to account for within-family relatedness. Pairwise group comparisons were subsequently evaluated using estimated marginal means (emmeans) package in R, which uses Tukey-adjusted p-values for multiple testing correction.

### Permutation analysis with rare and deleterious variants

We sought to evaluate whether rare variants, particularly those predicted to be deleterious, segregate within families more often than expected by chance, thereby contributing to disease burden in SCZ and BPD. Individual families were evaluated for rare deleterious variant segregation, defined as a rare deleterious variant for which all affected family members carried at least one copy of the variant and all unaffected family members were non-carriers. To examine the significance of such segregation within the complex family structures, a permutation analysis (n=10,000) was conducted separately for each segregating rare variant. More specifically, family affection status was randomly permuted while preserving the underlying genotype and LD structure, and segregation was re-evaluated across all families. This allowed us to calculate the probability of observing a higher number of families segregating each rare variant than expected by chance. To account for the polygenic nature of psychiatric disorders, variant-level permutation results were aggregated into gene-level and genome-wide burden analyses. To model the expected distribution of segregating events, a Poisson distribution was used for individual variant analyses, whereas one-sided empirical permutation tests were used for gene-level and genome-wide burden analyses.

### Gene-set based analyses using SAIGE-GENE+

We performed rare variant gene-set association analyses using SAIGE-GENE+ (35, 36). SAIGE implements a generalized linear mixed model framework to account for sample relatedness and population structure, while also improving statistical power in the presence of case–control imbalance. Gene sets were obtained from the C2 curated gene set collection of the Molecular Signatures Database (MSigDB), which contains 7,670 gene sets compiled from pathway databases and the biomedical literature (53). Prior to running SAIGE-GENE+, autosomal variants with a missing rate < 2% were retained for variance ratio estimation. From these variants, common variants (MAF ≥ 5%) were further LD pruned for generation of the principal component analysis (PCA) and genetic relationship matrix (GRM). For PCA construction, related individuals were excluded prior to principal component calculation, and the resulting principal components (PCs) were then projected onto all subjects. For construction of the C2 gene sets, only deleterious variants within pathway-associated genes, as identified by ANNOVAR annotations and CADD scores, were retained (n = 488,860). The first 4 PCs of the PCA were included in the model to account for population structure. To correct for multiple testing, we applied the False Discovery Rate (FDR) method (FDR-corrected p-value ≤ 0.05) (54).

## Results

### PRS of SCZ and BPD

We calculated the PRS (48) using our PIC dataset as the target data and the recent GWAS from PGC as the base dataset. Figure 1 shows the results of PRS analyses on both SCZ and BP cases versus population controls and cases versus unaffected relatives. The p-value threshold of 0.05 explained the greatest proportion of risk for SCZ (*R*^2^ = 0.246; p-value = 2.9×10^-5^) and BPD (*R*^2^ = 0.064; p-value = 9.6×10^-3^), respectively, when comparing cases versus population controls, and p-value threshold of 0.001 and 0.05 for SCZ (*R*^2^ = 0.003; p-value = 0.5) and BPD (*R*^2^ = 0.012; p-value = 0.2), respectively, for cases versus unaffected relatives analysis. Additional logistic regression analyses evaluating the association between PRS and disease status showed significant associations for SCZ (OR = 2.92, p = 3.05×10^-5^) and BPD (OR = 1.59, p = 9.03×10^-3^) when compared to population controls, indicating a strong genetic risk contribution. However, in family-based comparisons (cases versus unaffected relatives), the associations were weaker and not statistically significant (SCZ: OR = 1.13, p = 0.4; BPD: OR = 1.22, p = 0.2), likely due to shared genetic background. Overall, our PRS analysis yielded statistically significant results for both SCZ and BPD. The genetic variants included in the PRS, at specified p-value thresholds, explained a significant portion of the risk for these disorders. These results indicate that the identified genetic risk factors contribute meaningfully to SCZ and BPD susceptibility in our PIC population, and PRS effectively differentiates cases from general population controls.

**Figure 1.**
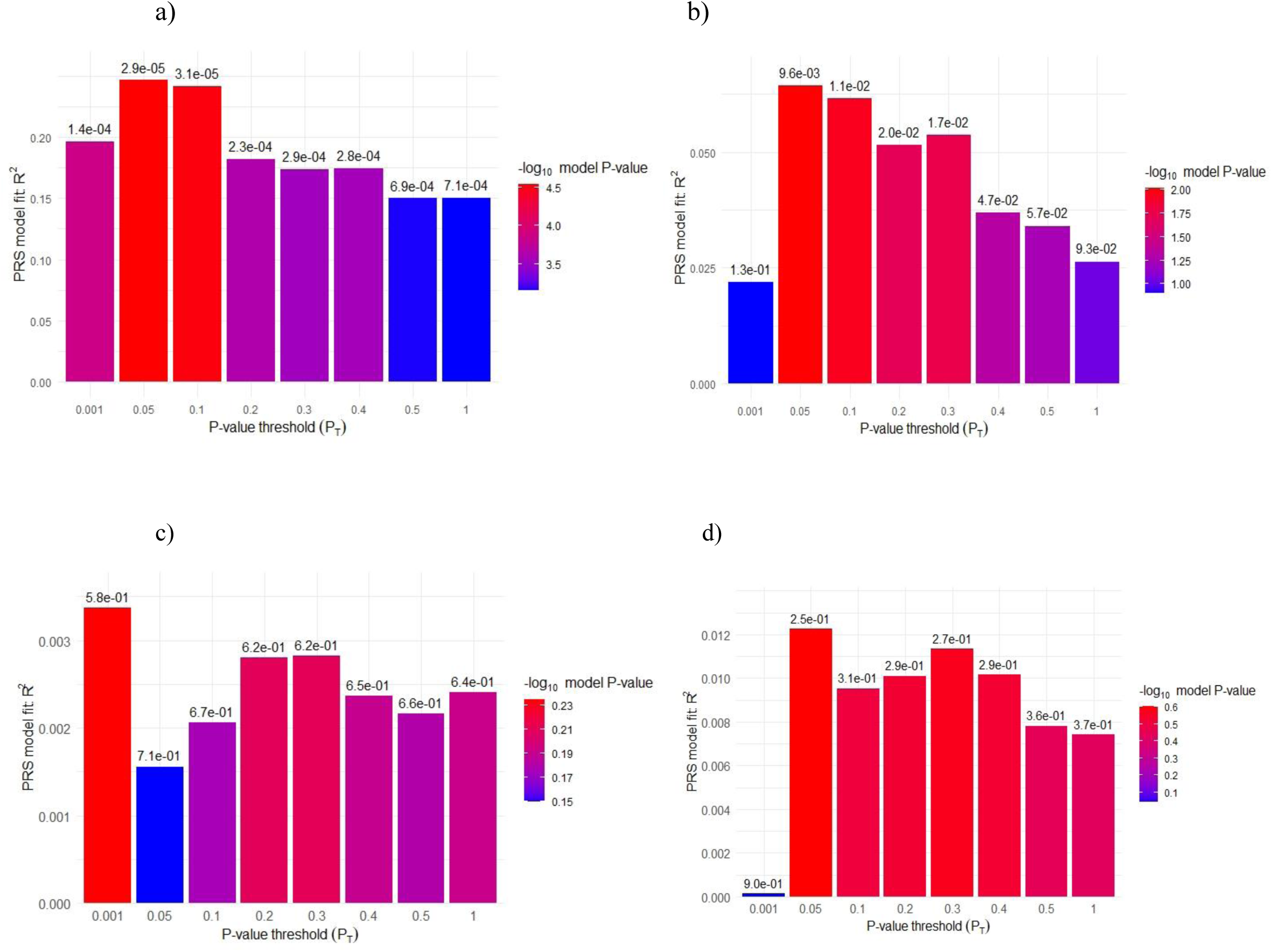
PRS analysis results for SCZ cases vs. population controls (a), BPD cases vs. population controls (b), SCZ cases vs. unaffected relatives (c), and BPD cases vs. unaffected relatives (d). The x-axis represents the p-value threshold (*P*_T_) used to select SNPs for inclusion in the PRS model. The y-axis shows the proportion of variance explained (*R*²) by the PRS in the phenotype, indicating the model fit at each *P*_T_.

### Burden analysis and permutation of rare deleterious variants

We used ANNOVAR for variant annotation and the CADD-PHRED score to identify rare deleterious variants. After filtering individuals based on excessive Mendelian errors, sex-check failures, and/or low-quality reads, 181 and 208 individuals for the SCZ and BPD analyses, respectively, were categorized into affected, unaffected, and control groups (Figures 2a and 2b). Figure 2 also depicts the rare and deleterious variant counts segregating in the controls, unaffected, and affected subjects. To ensure unbiased comparisons, the variant counts were adjusted for group size to account for differences in sample sizes. Based on previous studies (7, 16, 18, 55), we only focused on variant categories enriched in cases that were subsequently included in the ANNOVAR analysis.

**Figure 2.**
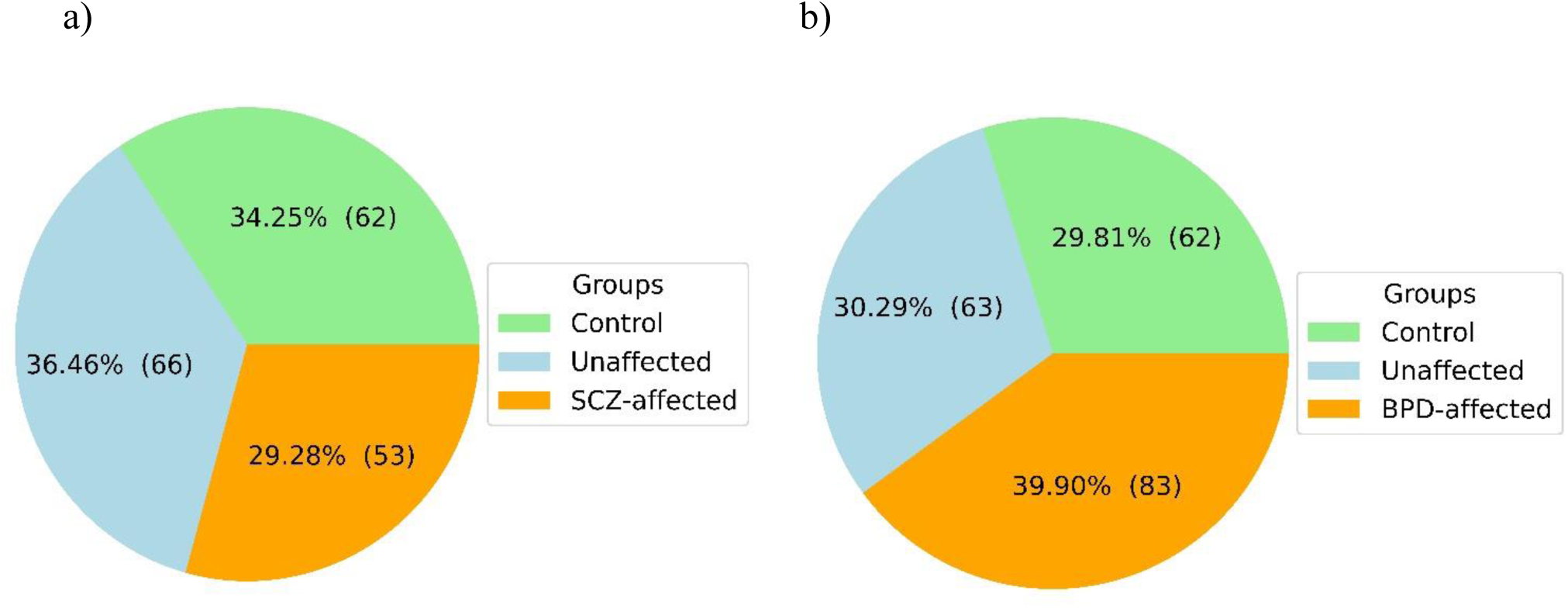
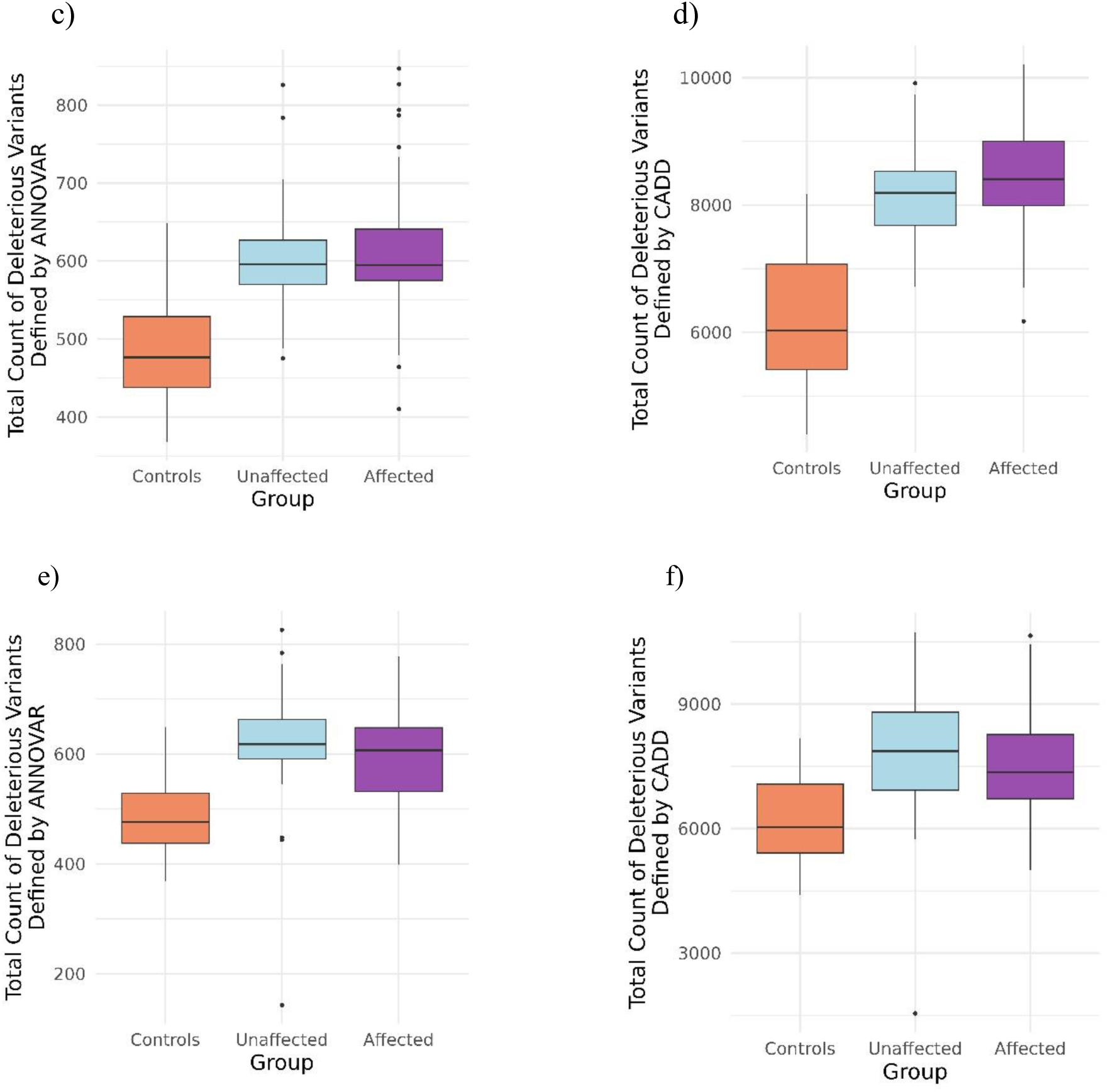
Status of the three groups for SCZ (a) and BPD (b), and total count of deleterious variants defined by ANNOVAR and CADD for SCZ (c and d) and BPD (e and f).

To assess group differences in the burden of deleterious variants while accounting for familial relatedness across all groups (affected vs. unaffected, affected vs. controls, controls vs. unaffected), linear mixed-effects models including family as a random effect were performed separately for both SCZ and BPD using deleteriousness defined by both ANNOVAR and CADD scores. Pairwise comparisons based on estimated marginal means with Tukey correction demonstrated significant differences between affected individuals and controls, as well as between controls and unaffected individuals, across both disorders (Figure 2). For ANNOVAR-defined deleterious variants, significant differences were observed between SCZ / BPD affected and controls (p < .0001) and between controls and SCZ / BPD unaffected individuals (p < .0001). Similar patterns were observed using CADD-defined deleterious variants, with significant differences between SCZ / BPD affected and controls (p < .0001) and between controls and SCZ / BPD unaffected individuals (p < .0001). No significant differences were observed between affected and unaffected individuals for either SCZ (ANNOVAR: p = 0.9922; CADD: p = 0.7642) or BPD (ANNOVAR: p = 0.5285; CADD: p = 0.6242). The lack of significant differences between affected and unaffected individuals likely reflects their shared familial and genetic background, as both groups originate from the same pedigrees and therefore carry overlapping genetic risk. In contrast, the observed significant differences involving population controls (i.e., affected vs. controls and unaffected vs. controls) are consistent with the genetic distinctiveness of the control group, which lacks the familial risk present in both affected individuals and their unaffected relatives. These findings suggest that unaffected family members may harbor an elevated burden of deleterious variants similar to affected individuals, potentially representing subclinical genetic susceptibility that does not manifest as disease and well-documented in multiplex families.

Permutation-based segregation analysis was performed to evaluate whether rare deleterious variants segregated within multiplex SCZ and BPD families more frequently than expected by chance. At the variant and gene levels, the SCZ analysis identified 4,745 variants and 415 genes with nominal significance (p ≤ 0.05), whereas the BPD analysis identified 9,215 variants and 564 genes showing nominal evidence of segregation (Supplementary Files 4-1 to 4-4). However, none of these associations survived FDR correction. At the genome-wide level, no significant enrichment of segregating rare deleterious variants was observed in either disorder. In SCZ, 710,231 observed segregating events were identified among 2,501,666 tested variants (permutation p = 1.0), while in BPD, 516,889 segregating events were identified among 2,091,460 tested variants (permutation p = 1.0). These findings suggest that although numerous variants demonstrated nominal segregation within families, the overall genome-wide burden of segregating rare deleterious variants did not exceed chance expectation under permutation testing, likely reflecting limited statistical power and the substantial multiple-testing burden inherent to genome-wide rare variant analyses in multiplex pedigrees.

### Gene-set based analyses

Gene-set–based rare variant association analyses were performed using SAIGE-GENE+, accounting for population structure, relatedness, and case–control imbalance. This framework is particularly well-suited for rare variant analyses, enabling robust detection of genetic associations while controlling for potential confounding factors. By leveraging SAIGE-GENE+, we aimed to identify biologically relevant gene sets associated with SCZ and BPD, while maintaining statistical rigor in the context of family-based WGS data. Analyses were conducted comparing SCZ and BPD cases to population controls using CADD ≥ 10 thresholds. SAIGE-GENE+ identified 483 and 583 nominally significant gene sets (p-value ≤ 0.05) for SCZ and BPD, respectively; however, none remained significant after multiple testing correction (FDR > 0.05) (Supplementary Files 5-1 to 5-4). Tables 1 and 2 summarize the gene sets strongly implicated in the biological mechanisms underlying SCZ and BPD, respectively.

**Table 1.** Gene sets biologically relevant to SCZ.

| Gene set | p-value | Beta | SE | FDR |
| --- | --- | --- | --- | --- |
| WP_OLIGODENDROCYTE_DEVELOPMENT | 0.000791507 | 5.964174056 | 1.777300982 | 0.502522038 |
| WP_AMPACTIVATED_PROTEIN_KINASE_SIGNALING | 0.002847193 | 14.02433302 | 4.700203439 | 0.502522038 |
| CHESLER_BRAIN_QTL_CIS | 0.003726882 | 5.807583667 | 2.002339748 | 0.508529301 |
| REACTOME_DOPAMINE_NEUROTRANSMITTER_RELEASE_CYCLE | 0.004371384 | 0.511908241 | 0.179614455 | 0.51074466 |
| REACTOME_SEROTONIN_NEUROTRANSMITTER_RELEASE_CYCLE | 0.004371384 | 0.511908241 | 0.179614455 | 0.51074466 |
| MEISSNER_ES_ICP_WITH_H3K4ME3_AND_H3K27ME3 | 0.005070578 | 0.382818917 | 0.136598322 | 0.51074466 |
| REACTOME_SEROTONIN_RECEPTORS | 0.005559498 | 0.237085664 | 0.085507442 | 0.51074466 |
| WP_WNT_SIGNALING_NETPATH | 0.007465453 | 2.939970938 | 1.098916782 | 0.538113132 |
| HOUSTIS_ROS | 0.007744105 | 0.52447066 | 0.196945255 | 0.538113132 |
| WONG_MITOCHONDRIA_GENE_MODULE | 0.008519815 | 467.9637464 | 177.8826522 | 0.538113132 |
| REACTOME_PROTEIN_PROTEIN_INTERACTIONS_AT_SYNAPSES | 0.009416577 | 461.8078293 | 177.8545385 | 0.538113132 |
| WP_NEUROINFLAMMATION_AND_GLUTAMATERGIC_SIGNALING | 0.009834879 | 115.2160057 | 44.63001059 | 0.538983624 |
| WP_GABA_AND_GLUTAMATE_SIGNALLING_IN_EPILEPTOGENESIS | 0.012603566 | 46.8056807 | 18.76145505 | 0.574454683 |
| REACTOME_ACETYLCHOLINE_NEUROTRANSMITTER_RELEASE_CYCLE | 0.01318028 | 0.368581219 | 0.148689771 | 0.574454683 |
| KEGG_PRIMARY_IMMUNODEFICIENCY | 0.013382324 | 0.591441555 | 0.239118024 | 0.574454683 |
| REACTOME_OTHER_INTERLEUKIN_SIGNALING | 0.013764317 | 0.720913391 | 0.292654974 | 0.574454683 |
| WP_MONOAMINE_TRANSPORT | 0.014453494 | 0.880069273 | 0.359830189 | 0.574454683 |
| JAIN_NFKB_SIGNALING | 0.015396698 | 11.72898101 | 4.840863296 | 0.574454683 |
| LEIN_MIDBRAIN_MARKERS | 0.021683497 | 408.0253443 | 177.7216956 | 0.595344249 |
| MIKKELSEN_ES_ICP_WITH_H3K4ME3_AND_H3K27ME3 | 0.0268257 | 638.2001773 | 288.2508011 | 0.628401315 |
| REACTOME_NOREPINEPHRINE_NEUROTRANSMITTER_RELEASE_CYCLE | 0.030314829 | 0.424335247 | 0.195911627 | 0.643348052 |
| REACTOME_GLUTAMATE_NEUROTRANSMITTER_RELEASE_CYCLE | 0.03286948 | 0.508388761 | 0.238269188 | 0.644089451 |
| WP_SYNAPTIC_SIGNALING_ASSOCIATED_WITH_AUTISM_SPECTRUM_DISORDER | 0.03667914 | 2.288190845 | 1.095185726 | 0.661205764 |
| REACTOME_NEUROTRANSMITTER_RELEASE_CYCLE | 0.040697324 | 1.414385547 | 0.691090191 | 0.663272531 |
| LEE_AGING_NEOCORTEX_DN | 0.04113755 | 4.611033082 | 2.257940642 | 0.663272531 |
| MEISSNER_BRAIN_HCP_WITH_H3K4ME2 | 0.043545683 | 0.298390823 | 0.147832556 | 0.672045109 |
| WP_OXIDATIVE_STRESS_RESPONSE | 0.044011567 | 0.651078541 | 0.323279445 | 0.672045109 |
| KEGG_NEUROTROPHIN_SIGNALING_PATHWAY | 0.044235913 | -24.82385733 | 12.33883056 | 0.672045109 |
| REACTOME_TCF_DEPENDENT_SIGNALING_IN_RESPONSE_TO_WNT | 0.048437629 | 349.83973 | 177.2678889 | 0.678999017 |

**Table 2.** Gene sets biologically relevant to BPD.

| Gene set | p-value | Beta | SE | FDR |
| --- | --- | --- | --- | --- |
| WP_CYTOKINECYTOKINE_RECEPTOR_INTERACTION | 4.34E-05 | 625.2648895 | 152.9296179 | 0.145137271 |
| REACTOME_MITOCHONDRIAL_PROTEIN_DEGRADATION | 0.001239096 | 3.773213851 | 1.168277351 | 0.517692925 |
| WP_AKT_SIGNALING_AND_ARTD_FAMILY_MEMBERS | 0.002915655 | 1.61355196 | 0.542098315 | 0.527024391 |
| WP_GLUCOCORTICOID_RECEPTOR_PATHWAY | 0.004257505 | 1.418718457 | 0.496328836 | 0.544762207 |
| REACTOME_ATP_DEPENDENT_CHROMATIN_REMODELERS | 0.00571148 | 0.474646269 | 0.171730536 | 0.544762207 |
| MIKKELSEN_MEF_ICP_WITH_H3K4ME3_AND_H3K27ME3 | 0.006783414 | 0.283735118 | 0.104803952 | 0.544762207 |
| KEGG_TRYPTOPHAN_METABOLISM | 0.008066717 | 1.104316622 | 0.416838948 | 0.544762207 |
| MARTIN_NFKB_TARGETS_UP | 0.008214771 | 0.527603935 | 0.199614627 | 0.544762207 |
| WP_GABA_AND GLUTAMATE_SIGNALLING_IN_EPILEPTOGENESIS | 0.008342383 | 59.29715372 | 22.47901579 | 0.544762207 |
| WU_ALZHEIMER_DISEASE_DN | 0.008594425 | 0.697170288 | 0.265307775 | 0.544762207 |
| WP_OPIOID_RECEPTOR_PATHWAYS | 0.011094275 | 0.59231816 | 0.233222271 | 0.544762207 |
| KEGG_STEROID_HORMONE_BIOSYNTHESIS | 0.01131673 | 4.141402771 | 1.635132098 | 0.544762207 |
| REACTOME_INTERLEUKIN_1_SIGNALING | 0.012087705 | -1.605829774 | 0.63988154 | 0.544762207 |
| REACTOME_SIGNALING_BY_WNT | 0.012138773 | 1793.41386 | 715.0532872 | 0.544762207 |
| WP_TCA_CYCLE_AKA_KREBS_OR_CITRIC_ACID_CYCLE | 0.013503545 | 0.295577946 | 0.119657228 | 0.544762207 |
| REACTOME_CELL_JUNCTION_ORGANIZATION | 0.014224771 | 990.0443692 | 403.8464522 | 0.544762207 |
| REACTOME_CELL_EXTRACELLULAR_MATRIX_INTERACTIONS | 0.014258335 | 0.362921768 | 0.148089728 | 0.544762207 |
| WP_SYNAPTIC_SIGNALING_ASSOCIATED_WITH_AUTISM_SPECTRUM_DISORDER | 0.015148428 | 1.100672697 | 0.453173472 | 0.550188174 |
| BIOCARTA_REELIN_PATHWAY | 0.016459871 | 0.273220093 | 0.113910254 | 0.558522371 |
| WP_WNT_SIGNALING_NETPATH | 0.016865785 | 0.839186999 | 0.351180015 | 0.558522371 |
| REACTOME_TNF_SIGNALING | 0.019845091 | 1.643728093 | 0.705685653 | 0.567637984 |
| KEGG_CYTOKINE_CYTOKINE_RECEPTOR_INTERACTION | 0.019883865 | 846.6014145 | 363.5772962 | 0.567637984 |
| DER_IFN_GAMMA_RESPONSE_UP | 0.021401107 | 0.518140533 | 0.225196991 | 0.574821688 |
| WP_MECP2_AND_ASSOCIATED_RETT_SYNDROME | 0.022187475 | 4.003460116 | 1.750419645 | 0.577814873 |
| REACTOME_DCC_MEDIATED_ATTRACTIVE_SIGNALING | 0.023442958 | 0.19702319 | 0.086942288 | 0.582918106 |
| BLALOCK_ALZHEIMERS_DISEASE_INCIPIENT_DN | 0.02515626 | 107.3805978 | 47.95929049 | 0.584629559 |
| MIKKELSEN_IPS_LCP_WITH_H3K27ME3 | 0.027357312 | 0.153959094 | 0.069779006 | 0.584629559 |
| WP_NOTCH_SIGNALING_WP61 | 0.028619717 | 4.33774834 | 1.981896719 | 0.58872442 |
| WP_PARKINSONS_DISEASE_PATHWAY | 0.032069784 | -0.420826846 | 0.196323324 | 0.598900087 |
| KEGG_MTOR_SIGNALING_PATHWAY | 0.032258361 | 0.635734238 | 0.296906355 | 0.598900087 |
| WP_HISTONE_MODIFICATIONS | 0.037791956 | 5.01383484 | 2.413858288 | 0.598900087 |
| REACTOME_PI3K_AKT_SIGNALING_IN_CANCER | 0.03832887 | 65.54907521 | 31.64603463 | 0.598900087 |
| DIERICK_SEROTONIN_FUNCTION_GENES | 0.038719816 | -0.21112374 | 0.102132654 | 0.598900087 |
| WP_CIRCADIAN_RHYTHM_GENES | 0.040717013 | 3168.315509 | 1548.238472 | 0.598900087 |
| REACTOME_LONG_TERM_POTENTIATION | 0.04229029 | -0.206875453 | 0.10187636 | 0.606388776 |
| REACTOME_CREB1_PHOSPHORYLATION_THROUGH_NMDA_RECEPTOR_MEDIATED_ACTIVATION_OF_RAS_SIGNALING | 0.043531616 | -0.272029079 | 0.13476306 | 0.606388776 |
| PID_PI3KCI_AKT_PATHWAY | 0.043573202 | 0.577717369 | 0.286257543 | 0.606388776 |
| REACTOME_UNBLOCKING_OF_NMDA_RECEPTORS GLUTAMATE_BINDING_ANDACTIVATION | 0.0436205 | -0.235501166 | 0.116716513 | 0.606388776 |
| REACTOME_PRC2_METHYLATES_HISTONES_AND_DNA | 0.048717079 | 0.150173753 | 0.076189358 | 0.60834285 |
| WP_NEURODEGENERATION_WITH_BRAIN_IRON_ACCUMULATION_NBIA_SUBTYPE S_PATHWAY | 0.049207072 | 0.515384996 | 0.262043326 | 0.60834285 |

SAIGE-GENE+ results using C2 curated gene sets revealed multiple biologically relevant pathways implicating established mechanisms underlying SCZ. A prominent signal was observed in gene sets related to neurotransmission and synaptic function, encompassing dopaminergic, serotonergic, glutamatergic, and monoaminergic signaling systems and neurotransmitter transporters. In addition to neurotransmission, multiple enriched gene sets mapped to synaptic structure and neuronal communication pathways, indicating disruption of synaptic integrity and signaling. Gene sets associated with neurodevelopmental processes and brain-specific regulatory programs were also enriched, including pathways involving oligodendrocyte biology, myelination, and cortical development. A substantial proportion of enriched gene sets involved immune and inflammatory signaling pathways, including cytokine signaling, interleukin pathways, and NF-κB–related processes. Additional enriched relevant gene sets included Wnt signaling, neurotrophin signaling and MAPK/AMPK-related energy regulation pathways, epigenetic regulation, and oxidative stress and mitochondrial function pathways.

In BPD, notably, several gene sets directly related to synaptic neurotransmission and plasticity were identified, including glutamatergic neurotransmission and NMDA receptor signaling. In addition, gene sets associated with neurodevelopmental processes and synaptic organization were observed. Gene sets linked to neurodegeneration and brain disorders, including those implicated in Alzheimer’s and Parkinson’s disease, were also enriched. Furthermore, several gene sets related to neuroendocrine regulation and behavior-related signaling, including glucocorticoid signaling and circadian rhythm processes, were identified. Gene sets related to serotonin metabolism were also present. Beyond directly brain-related gene sets, enrichment was observed in biological systems increasingly recognized in psychiatric disorders, including immune and inflammatory signaling, mitochondrial and metabolic function, and epigenetic regulation. Additionally, enrichment of signaling pathways such as Wnt and Akt/mTOR were also observed. While the majority of gene sets were not specific to psychiatric phenotypes, the suggestive enrichment of core neurobiological pathways supports the relevance of our findings to SCZ and BPD-related mechanisms.

### Cross-disorder gene sets overlap

Among the most relevant gene sets identified in SCZ (n = 29) and BPD (n = 40), comparison of nominally significant gene sets revealed limited direct overlap at the individual gene-set level, with only three pathways shared between the two analyses: GABA and glutamate signaling, synaptic signaling, and Wnt signaling. Despite this limited overlap, broader functional convergence was observed across key biological domains. Both disorders showed nominal associations with pathways related to neurotransmission and synaptic signaling, including glutamatergic and GABAergic processes, as well as monoaminergic systems in SCZ (dopamine, serotonin, acetylcholine, and norepinephrine release cycles). Immune and inflammatory pathways were also represented in both disorders, including cytokine signaling and NF-κB–related processes. Additional convergence was observed in gene sets related to Wnt signaling, epigenetic regulation (including histone modification and chromatin remodeling), and mitochondrial and metabolic function, including oxidative stress–related processes. These findings indicate that, although overlap at the individual gene-set level is limited, SCZ and BPD share convergent biological mechanisms at the pathway level. Figure 3 demonstrates the distribution of nominally associated gene sets across major biological categories identified in the SCZ and BPD analyses. The largest proportion of gene sets was related to neurotransmission and synaptic function, followed by neurodevelopmental, immune/inflammatory, and major signaling pathways, supporting the involvement of multiple convergent neurobiological processes in psychiatric disease susceptibility (Supplementary File 6).

**Figure 3.**
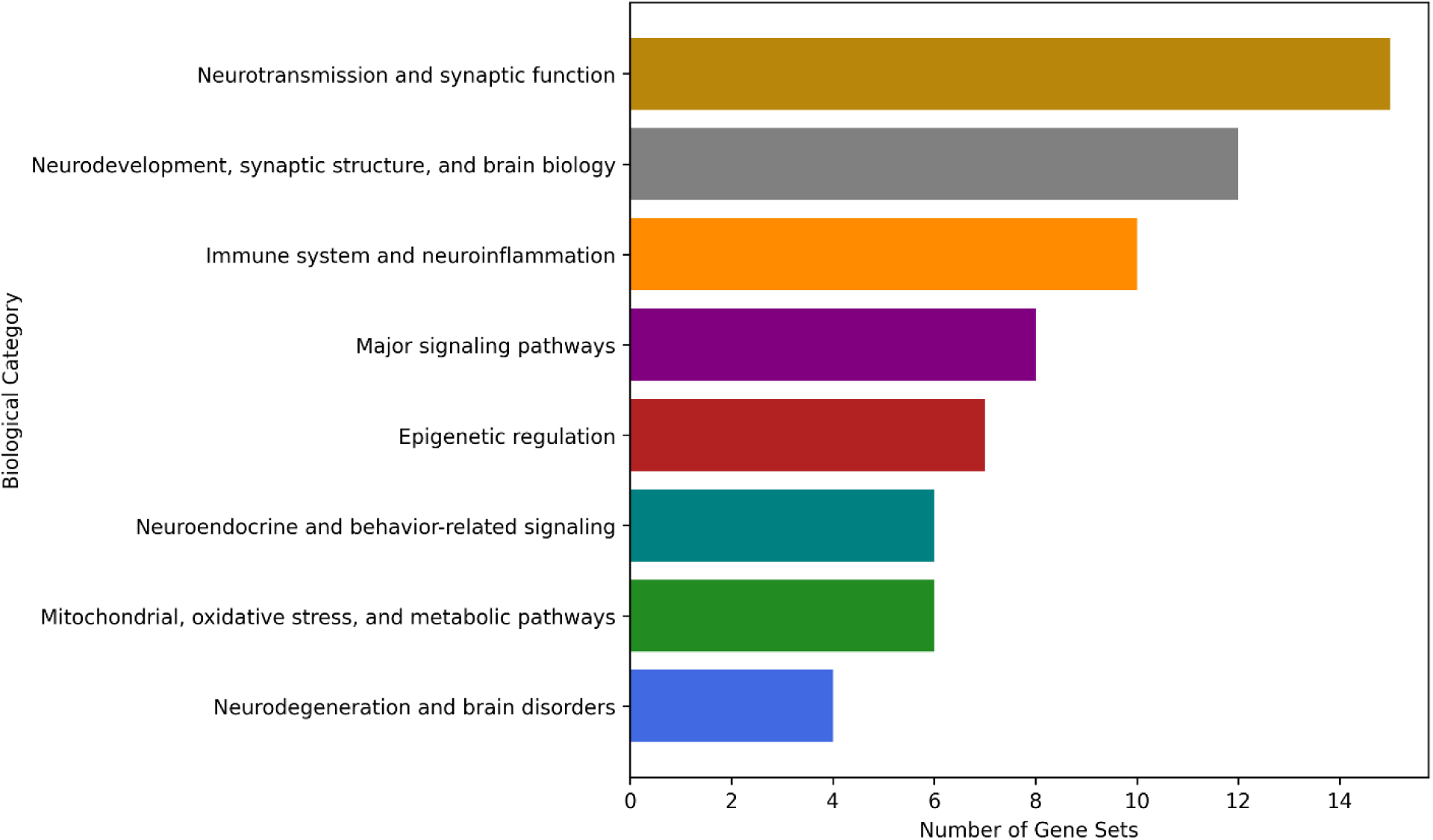
Distribution of nominally associated gene sets across major biological categories identified in SAIGE-GENE+ analyses in SCZ and BPD.

## Discussion

In this study, we investigated unique multiplex families from the PIC for evidence of segregating rare variants contributing to the genetic basis of SCZ and BPD. Using gene-set based rare variant association analysis on WGS data, our analyses revealed suggestive associations between gene sets harboring rare variants in both disorders, particularly in gene sets involved in neurotransmission, synaptic function and structure, neurodevelopment, and neuroinflammation as well as major signaling pathways. These findings are consistent with previous reports and provide compelling evidence for the involvement of these gene sets in the pathophysiology of SCZ and BPD, as well as offer new insights into the shared genetic architecture of these complex neuropsychiatric disorders.

### Neurotransmission and synaptic function

Neurotransmitter systems play an important role in the pathophysiology of neuropsychiatric disorders (56), and dysregulation at different levels of these systems, particularly, dopaminergic and glutamatergic neurotransmitter systems, likely contributes to manic symptoms and impaired synaptic transmission/neuroplasticity observed in BPD and other neuropsychiatric disorders (57, 58). The dopamine system closely interacts with other brain systems including the glutamate system, and disruptions in this interplay (dopaminergic/glutamatergic system) likely contribute to positive, negative, and cognitive symptoms of SCZ, underscoring its role in the disorder’s pathophysiology (59). Furthermore, there is substantial evidence linking presynaptic and postsynaptic dopaminergic neuron abnormalities to mental illness onset (60, 61). Consistent with this, rare variant association studies in SCZ cases have previously identified risk genes involved in the regulation of the neurotransmitter transport pathway, which supported the involvement of neural system perturbations in SCZ (62). In our study, gene sets such as dopamine neurotransmitter, serotonin neurotransmitter, and glutamate neurotransmitter which are involved in various aspects of neurodevelopment and neurotransmission processes, were suggestively associated with SCZ, and our results align with previous studies linking dopaminergic dysfunction to SCZ and BPD (60, 61, 63). Further evidence from clinical studies and postmortem brain analyses indicates N-methyl-D-aspartate receptor (NMDAR) hypofunction in SCZ. Studies have identified common and rare genetic variants enriched in glutamatergic postsynaptic proteins, including components of the NMDAR complex, as well as the postsynaptic density, that are associated with increased risk of SCZ (14, 16, 27). Dysfunction of the NMDAR-associated postsynaptic signaling complex, which is critical for synaptic plasticity and cognitive processes, has therefore been proposed as a key contributor to SCZ pathogenesis (64). In addition to dopaminergic and glutamatergic systems, alterations in serotonergic signaling are implicated across multiple psychiatric disorders ranging from anxiety to SCZ (65). In SCZ, serotonergic dysfunction was associated with negative symptoms, suggesting that regulation of serotonin release may offer therapeutic benefit, particularly in deficit subtypes, underscoring the importance of serotonin signaling in the disorder (66). Previous studies also link presynaptic dysfunction to psychiatric disorders, with rare variants in genes crucial for neurotransmitter release at the presynaptic terminal, implicated in the pathogenesis of SCZ and BPD (31, 67). These observations are consistent with our findings including suggestive associations with gene sets such as glutamate neurotransmitter and serotonin receptors. Our analysis also identified suggestive associations of several gene sets mostly related to synaptic function and neurotransmission including GABA and glutamate signaling in both SCZ and BPD, as well as long-term potentiation and NMDA receptors and glutamate binding with BPD. These findings support previous evidence implicating dysregulation of synaptic vesicle function (GABA-enriched and Glu-enriched), particularly in GABAergic and glutamatergic systems in both SCZ and BPD (57, 63). Disruptions in GABA/GABAergic signaling and the excitatory/inhibitory neurotransmission balance may contribute to the development of several psychiatric disorders including SCZ and BPD (68–70). Long-term potentiation (LTP) has a crucial role in synaptic plasticity, a process critical for learning and memory, which is impaired in SCZ and BPD, highlighting its involvement in synaptic dysfunction (63, 71, 72). In addition, dysregulation of NMDAR signaling and decreased expression of gene networks that are enriched for synaptic functions in cerebellar neurons, have been implicated in BPD pathophysiology (73).

### Neurodevelopment and synaptic structure

The synaptic signaling gene set which is involved in synaptic dysfunction (63, 71), showed associations with both SCZ and BPD in our study. This finding is consistent with prior WGS studies of multiplex families, which have revealed de-novo rare/damaging variants in genes involved in neurodevelopmental pathways including cranial nerve development/morphogenesis, neuron projection extension/morphogenesis (26), and synaptic function and localization (18). Synaptic signaling is central to SCZ and BPD pathophysiology, with converging evidence indicating that disrupted synaptic development and function contribute to impaired neural connectivity and the emergence of core clinical symptoms (14, 74). Multiple factors including changes in synaptic architecture, dysregulated expression of plasticity-related genes, and abnormal neurotransmission (dopaminergic, serotonergic, and glutamatergic) likely contribute to these deficits, affecting synaptic plasticity, neural communication, and cognition (75). In addition, several gene sets related to neurodevelopment and brain structure were suggestively associated with SCZ and BPD, including oligodendrocyte development, midbrain markers, brain HCP with H3K4ME2 in SCZ, and Reelin, MECP2, and Cell Extracellular Matrix Interaction in BPD. Prior brain studies have reported reduced expression of genes involved in oligodendrocyte function and myelination in both SCZ and BPD, suggesting that myelin and oligodendrocyte abnormalities may contribute to these disorders by disrupting signal transmission, altering oligodendrocyte distribution and function, along with disrupted myelination patterns, with evidence from postmortem studies showing altered myelin-related gene expression (76–78). These abnormalities may also underlie broader neurobiological deficits, potentially interacting with dopaminergic neurotransmission systems implicated in the disorders (78). Studies also reported a higher prevalence of midline brain abnormalities across mood and psychotic disorders, including failure of the septum pellucidum to fuse and absence of the adhesio interthalamica (79). In parallel, epigenetic dysregulation such as dysregulated H3K4 methylation has been implicated in psychiatric disorders by acting as a molecular link between environmental influences and neuronal and glial genomic regulation in the brain (80). Consistent with these findings, gene sets such as Reelin and MECP2 further highlight the importance of neurodevelopmental and synaptic regulatory mechanisms. Reelin is an extracellular matrix glycoprotein that regulates neuronal migration and synaptic plasticity, enhancing glutamatergic and GABAergic signaling and promoting synaptic maturation, and its loss or disrupted signaling is linked to neurodevelopmental abnormalities and psychiatric disorders (81). Also, Methyl-CpG–binding protein 2 (MeCP2) is a highly expressed brain transcriptional regulator that binds methylated DNA to control neuronal development and synaptic plasticity, and its alterations are also implicated in stress-related conditions such as major depression (82). Additionally, the extracellular matrix (ECM) plays key roles in neurodevelopment by regulating neural progenitor proliferation and differentiation, as well as shaping neuronal morphology such as axonal and dendritic elongation regulating their connectivity and cortical organization (83).

### Immune system and neuroinflammation

Our analysis identified suggestive associations with multiple immune and inflammation-related gene sets, including neuroinflammation and glutamatergic signaling, immunodeficiency, interleukin signaling, NF-kB signaling in SCZ, and cytokine-cytokine receptor interaction, interleukin-1 signaling, and NF-kB targets in BPD. These findings are consistent with growing evidence implicating immune dysregulation and neuroinflammatory processes in the pathophysiology of both disorders. SCZ involves widespread immune dysregulation, spanning innate and adaptive, humoral and cellular systems, with inflammatory activation observed in a substantial subset of patients. Peripheral immune changes may drive neuroinflammation, contributing to cognitive deficits, brain alterations, and disease pathogenesis (84). Notably, neuroinflammation especially in the dorsolateral prefrontal cortex, has been observed in a subset of individuals with SCZ and is marked by elevated levels of inflammatory markers, including multiple cytokines, and also dysregulation in chemokine expression and Wnt/β-catenin pathway (85–87). Similarly, immune dysfunction has also been implicated in cognitive impairment in BPD, with neuroinflammation potentially disrupting brain regions involved in memory and learning (88). Low-grade peripheral inflammation may trigger neuroinflammatory cascades in the central nervous system (CNS), impairing neuronal homeostasis and synaptic function (89). Elevated inflammatory markers are associated with psychiatric symptoms across disorders, with serum IL-1β and IL-16 correlating with symptom severity in SCZ, and increased IL-1β levels observed in adolescents with BPD (90, 91). In addition, higher NF-kB signaling pathway activity, reflected by elevated transcript levels for NF-kB family members, has been linked to increased cortical immune activation in SCZ and has also been reported in BPD, further supporting a shared role of inflammatory signaling mechanisms across these disorders (91, 92).

### Neuroendocrine and serotonin signaling

We found suggestive associations with several neuroendocrine and signaling-related gene sets in BPD including opioid receptor pathways, glucocorticoid receptor pathway, circadian rhythm genes, and serotonin function genes, supporting the involvement of stress-response systems and neuromodulatory signaling in the pathophysiology of BPD. In BPD, studies reported increased μ-opioid receptor (MOR) expression, suggesting dysregulation of the endogenous opioid system, with MOR overexpression linked to manic symptoms (93). Additional evidence implicates also altered kappa-opioid receptor (OPRK1) signaling in mood disorder pathophysiology (94). BPD is characterized by hypothalamic pituitary adrenal (HPA) axis hyperactivity, glucocorticoid insensitivity, and altered serotonin and inflammatory signaling, and dysregulation of intracellular pathways particularly glucocorticoid receptor (GR) may contribute to its pathophysiology (95, 96). HPA axis dysfunction in BPD may contribute to treatment resistance, relapse, poorer outcomes, and cognitive deficits, due to the disruption of the balance between glucocorticoid receptor and mineralocorticoid receptor function, further disrupting normal stress hormone regulation, and causing imbalance HPA axis activity (96). Further evidence also show that disrupted sleep and circadian rhythms are linked to clinical symptoms of mood disorders such as BPD. Both HPA axis GR pathway play key roles in regulating circadian rhythms, and reduced circadian rhythmicity has been shown to distinguish bipolar from unipolar disorder, and is associated with greater symptom severity and longer illness duration in BPD patients (95, 97). In parallel, serotonergic dysfunction has been implicated in mood regulation in BPD, with evidence of reduced central serotonergic activity during BPD depressive episodes (98). Altered expression of serotonin receptors including 5-HT1A, 5-HT1B, and 5-HT2A across mood disorders and SCZ further supports serotonergic signaling dysregulation in psychiatric disorders (99).

### Major signaling pathways

We observed enrichment of multiple core signaling pathways, including Wnt signaling in both SCZ and BPD, neurotrophin/MAPK and AMPK signaling in SCZ, and the PI3K/Akt pathway in BPD. These pathways play central roles in neurodevelopment, synaptic plasticity, and cellular homeostasis, highlighting their relevance to psychiatric disease biology. The Wnt signaling pathway is critical for neurodevelopment and regulating the function and structure of the adult nervous system, and its dysregulation, reflected in altered gene expression and protein levels, has been implicated in both SCZ and BPD (100, 101). Its interactions with neurotransmitter systems (i. e. dopamine, glutamate, GABA, serotonin) and neuroprotective roles further highlight its relevance to disease pathophysiology (102). In BPD, postmortem analyses have identified abnormalities in the Wnt/GSK3 signaling pathway within brain regions involved in mood regulation (103). Neurotrophins are key regulators of neurodevelopment and adult synaptic connectivity in the brain, and disruption of neurotrophin signaling pathways particularly those involving brain-derived neurotrophic factor (BDNF) and downstream MAPK/ERK cascades, as well as genetic variation in these pathways have been implicated in psychiatric disorders including SCZ (104, 105). Similarly, disruption in AMPK signaling has also been linked to neuronal structural abnormalities, reduced lifespan, and increased risk of neuropsychiatric symptoms (106).

Furthermore, in BPD, dysfunction of intracellular signaling pathways such as PI3K/Akt/mTOR particularly reduced Akt–mTOR signaling in the prefrontal cortex further underscore the role of disrupted cellular signaling in disease pathophysiology (107, 108). Reduced Akt activity in this region has been shown to drive cognitive deficits alongside changes in synaptic connectivity and function (108).

### Mitochondrial pathways

Our results highlight the involvement of gene sets related to mitochondrial function and oxidative stress, including mitochondria gene module, oxidative stress response, and mitochondrial protein degradation in both SCZ and BPD. In SCZ, increased oxidative stress, characterized by elevated lipid damage and reduced antioxidant defenses, has been associated with symptom severity (109, 110). Mitochondrial dysfunction, including altered gene expression, affects neural development and neuronal functions, and likely interacts with neurotransmitter abnormalities, oxidative stress, and inflammation to drive disease pathology, with evidence implicating bidirectional links between mitochondrial reactive oxygen species (ROS) and pro-inflammatory cytokines observed in SCZ and BPD (111, 112). Similarly, BPD may involve altered mitochondrial structure and function, along with neuroimmune dysregulation, and disrupted metabolic and oxidative stress pathways (113). Together, these findings highlight mitochondrial dysfunction as a shared biological mechanism contributing to cellular and synaptic deficits in SCZ and BPD.

### Epigenetic regulation

We also detected the involvement of epigenetic regulatory pathways, including histone modification and H3K4ME3_AND_H3K27ME3 gene sets, in both SCZ and BPD. Epigenetic mechanisms, including DNA methylation and histone modifications, play central roles in regulating gene expression and key cellular processes (transcriptional activity, chromatin folding, cell division and apoptotic processes, and DNA damage and repair) in SCZ (114). In BPD, altered histone H3 acetylation and gene-specific hypermethylation in the frontal cortex highlight the link between disrupted epigenetic regulation and neuroinflammation, synaptic function, and neuroprotection (115). Overall, these results suggest that epigenetic dysregulation may serve as a key mechanism connecting genetic variation to altered gene expression and subsequent neuronal dysfunction in both SCZ and BPD.

Comparison of nominally significant gene sets in SCZ and BPD revealed some overlap at the individual pathway level. However, broader functional convergence was evident, with both disorders implicating neurotransmission and synaptic signaling (including glutamatergic and GABAergic pathways), immune and inflammatory processes, Wnt signaling, epigenetic regulation, and mitochondrial/metabolic functions. These findings suggest shared biological mechanisms despite minimal direct pathway overlap.

## Conclusion

In summary, our gene set-based rare variant analyses in multiplex families from the PIC revealed the contribution of rare genetic variation in several gene sets related to neurotransmission and synaptic function, neurodevelopment, immune and inflammatory processes, intracellular signaling pathways, mitochondrial function, and epigenetic regulation. Our results strengthen the hypothesis that dysregulation in neurodevelopmental, synaptic, and brain development pathways are at the core of the underlying pathogenesis of these psychiatric disorders. The observed convergence across pathways such as synaptic signaling, glutamatergic and GABAergic signaling, Wnt and PI3K/Akt signaling, neuroinflammation, and mitochondrial dysfunction underscores shared mechanisms underlying SCZ and BPD, despite their clinical heterogeneity. Importantly, our study highlights the value of gene set–based approaches for capturing the cumulative effects of rare variants and identifying biologically relevant pathways associated with SCZ and BPD. Consistent with prior genetic studies, our results further emphasize the highly polygenic nature underlying these phenotypes and support the integration of rare variant analyses into studies of complex neuropsychiatric disorders. By focusing on multiplex families from a genetically isolated population, our work demonstrates the power of family-based designs to detect both shared and disorder-specific genetic risk factors, particularly rare variants that segregate within families. This approach also contributes meaningfully to the advancement of precision psychiatry, especially for complex traits like SCZ and BPD. However, the identified gene sets and variants might have functional implications and should be evaluated in future studies across diverse ethnic backgrounds, potentially informing the development of targeted therapeutic strategies for the treatment of SCZ and BPD. Overall, our study underscores the importance of integrating rare variant, gene-set based, and family-based analyses to dissect the complex genetic architecture of SCZ and BPD.

## Data Availability

All data produced in the present study are available upon reasonable request to the authors

## Acknowledgments

This work was funded by VA grant I01CX001380.

