## Supplementary File 3 for "Gene-Set Based Rare Variant Association Analysis of Whole Genome Sequencing Data in the Portuguese Island Collection for Schizophrenia and Bipolar Disorder"

**MDS of PIC and Iberian-Spanish 1000 genome**

By including the Iberian-Spanish dataset, we provided a reference population to compare against the PIC population to investigate the genetic relationships and population structure of the PIC relative to a closely related yet distinct population. This comparison was essential to assess the genetic distinctiveness of the PIC population, which could have implications for studying disease associations or genetic traits within an isolated population.

Filtering parameters included MAF ≤ 0.05 and linkage disequilibrium (LD) pruning (using --indep-pairwise 5 50 0.3). For MDS analysis, pairwise Identity-by-State (IBS) distances (--cluster) and the first 20 dimensions of the MDS plot (---mds-plot 20) were used as parameters.

The MDS analysis showed that these two populations cluster separately, indicating distinct genetic differences between them (Supplementary Figure). The separate clustering of a few families within the PIC dataset including 8529, 8539, and 8540 (all from Madeira) suggests the presence of subpopulation structures or unique genetic backgrounds within the PIC population itself. This could be due to various factors such as genetic drift, founder effects, or recent population admixture. Therefore, by comparing the PIC population with a well-defined reference population, we highlighted the genetic differences between these two populations and confirmed the genetic distinctiveness of the PIC population.

**Supplementary Figure.** MDS plot of the PIC families compared to the Iberian-Spanish 1000 genome (1000G_IBS) population.

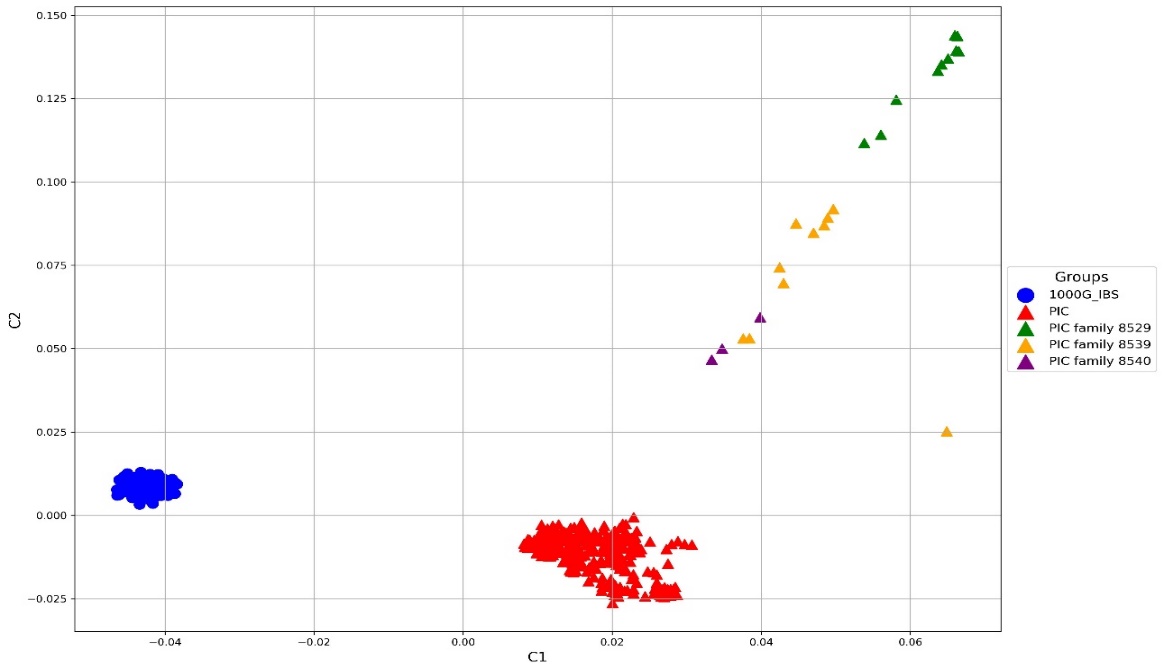
