## Supplementary File 6 for "Gene-Set Based Rare Variant Association Analysis of Whole Genome Sequencing Data in the Portuguese Island Collection for Schizophrenia and Bipolar Disorder"

Gene sets in each category in SAIGE-GENE+ SCZ analysis

| **Biological category** | **Gene sets** |
| --- | --- |
| Neurotransmission and synaptic function | • REACTOME_DOPAMINE_NEUROTRANSMITTER_RELEASE_CYCLE  • REACTOME_SEROTONIN_NEUROTRANSMITTER_RELEASE_CYCLE  • REACTOME_ACETYLCHOLINE_NEUROTRANSMITTER_RELEASE_CYCLE  • REACTOME_NOREPINEPHRINE_NEUROTRANSMITTER_RELEASE_CYCLE  • REACTOME_GLUTAMATE_NEUROTRANSMITTER_RELEASE_CYCLE  • REACTOME_NEUROTRANSMITTER_RELEASE_CYCLE  • REACTOME_SEROTONIN_RECEPTORS  • WP_MONOAMINE_TRANSPORT  • WP_GABA_AND_GLUTAMATE_SIGNALLING_IN_EPILEPTOGENESIS  • REACTOME_PROTEIN_PROTEIN_INTERACTIONS_AT_SYNAPSES  • WP_SYNAPTIC_SIGNALING_ASSOCIATED_WITH_AUTISM_SPECTRUM_DISORDER |
| Neurodevelopment, synaptic structure, and brain biology | • WP_OLIGODENDROCYTE_DEVELOPMENT  • LEIN_MIDBRAIN_MARKERS  • CHESLER_BRAIN_QTL_CIS  • MEISSNER_BRAIN_HCP_WITH_H3K4ME2  • LEE_AGING_NEOCORTEX_DN  • KEGG_NEUROTROPHIN_SIGNALING_PATHWAY |
| Mitochondrial / oxidative stress | • WONG_MITOCHONDRIA_GENE_MODULE  • WP_OXIDATIVE_STRESS_RESPONSE  • HOUSTIS_ROS  • WP_AMPACTIVATED_PROTEIN_KINASE_SIGNALING |
| Immune system and neuroinflammation | • WP_NEUROINFLAMMATION_AND_GLUTAMATERGIC_SIGNALING  • KEGG_PRIMARY_IMMUNODEFICIENCY  • REACTOME_OTHER_INTERLEUKIN_SIGNALING  • JAIN_NFKB_SIGNALING |
| Epigenetic regulation | • MEISSNER_ES_ICP_WITH_H3K4ME3_AND_H3K27ME3  • MIKKELSEN_ES_ICP_WITH_H3K4ME3_AND_H3K27ME3 |
| WNT / developmental signaling | • WP_WNT_SIGNALING_NETPATH  • REACTOME_TCF_DEPENDENT_SIGNALING_IN_RESPONSE_TO_WNT |

Gene sets in each category in SAIGE-GENE+ BPD analysis

| **Biological category** | **Gene sets** |
| --- | --- |
| Immune system and neuroinflammation | • WP_CYTOKINECYTOKINE_RECEPTOR_INTERACTION  • KEGG_CYTOKINE_CYTOKINE_RECEPTOR_INTERACTION  • REACTOME_INTERLEUKIN_1_SIGNALING  • REACTOME_TNF_SIGNALING  • MARTIN_NFKB_TARGETS_UP  • DER_IFN_GAMMA_RESPONSE_UP |
| Major signaling pathways | • REACTOME_SIGNALING_BY_WNT  • WP_WNT_SIGNALING_NETPATH  • KEGG_MTOR_SIGNALING_PATHWAY  • REACTOME_PI3K_AKT_SIGNALING_IN_CANCER  • WP_AKT_SIGNALING_AND_ARTD_FAMILY_MEMBERS  • PID_PI3KCI_AKT_PATHWAY |
| Neurodevelopment and synaptic structure | • WP_SYNAPTIC_SIGNALING_ASSOCIATED_WITH_AUTISM_SPECTRUM_DISORDER  • BIOCARTA_REELIN_PATHWAY  • WP_MECP2_AND_ASSOCIATED_RETT_SYNDROME  • REACTOME_DCC_MEDIATED_ATTRACTIVE_SIGNALING  • REACTOME_CELL_JUNCTION_ORGANIZATION  • REACTOME_CELL_EXTRACELLULAR_MATRIX_INTERACTIONS |
| Epigenetic regulation | • REACTOME_ATP_DEPENDENT_CHROMATIN_REMODELERS  • WP_HISTONE_MODIFICATIONS  • REACTOME_PRC2_METHYLATES_HISTONES_AND_DNA  • MIKKELSEN_IPS_LCP_WITH_H3K27ME3  • MIKKELSEN_MEF_ICP_WITH_H3K4ME3_AND_H3K27ME3 |
| Neuroendocrine / behavior-related signaling | • WP_OPIOID_RECEPTOR_PATHWAYS  • WP_GLUCOCORTICOID_RECEPTOR_PATHWAY  • KEGG_STEROID_HORMONE_BIOSYNTHESIS  • WP_CIRCADIAN_RHYTHM_GENES |
| Neurodegeneration / brain disorders | • WP_PARKINSONS_DISEASE_PATHWAY  • WU_ALZHEIMER_DISEASE_DN  • BLALOCK_ALZHEIMERS_DISEASE_INCIPIENT_DN  • WP_NEURODEGENERATION_WITH_BRAIN_IRON_ACCUMULATION_NBIA_SUBTYPES_PATHWAY |
| Mitochondrial / metabolic pathways | • REACTOME_MITOCHONDRIAL_PROTEIN_DEGRADATION  • WP_TCA_CYCLE_AKA_KREBS_OR_CITRIC_ACID_CYCLE |
| Neurotransmission and synaptic function | • WP_GABA_AND_GLUTAMATE_SIGNALLING_IN_EPILEPTOGENESIS  • REACTOME_LONG_TERM_POTENTIATION  • REACTOME_UNBLOCKING_OF_NMDA_RECEPTORS_GLUTAMATE_BINDING_ANDACTIVATION  • REACTOME_CREB1_PHOSPHORYLATION_THROUGH_NMDA_RECEPTOR_MEDIATED_ACTIVATION_OF_RAS_SIGNALING |
| Serotonin signaling | • DIERICK_SEROTONIN_FUNCTION_GENES  • KEGG_TRYPTOPHAN_METABOLISM |
